# The Neurocognitive Profile of Post-operative Paediatric Cerebellar Mutism Syndrome: A Systematic Review

**DOI:** 10.1101/2025.02.21.25322700

**Authors:** Bethany M. Horne, Annisha A. Attanayake, Kristian Aquilina, Tara Murphy, Charlotte P. Malcolm

**Author notes:** Correspondence to: Dr Charlotte Malcolm, Neuropsychology Service, Psychological and Mental Health Services, Great Ormond Street Hospital, WC1N 3JH, UK.

## Abstract

**AIM:** To systematically review neurocognitive outcomes associated with Post-operative Paediatric Cerebellar Mutism Syndrome (PPCMS), comparing children with and without PPCMS after posterior fossa tumour surgery, and in relation to moderating demographic and clinical risk factors. **METHODS** PsycInfo, Medline and Embase databases were systematically searched up to December 2024. Studies of children aged 2-18 years with PPCMS who had undergone standardised neurocognitive assessment were included. Quality was appraised using Institute of Health Economics Quality Appraisal Checklist for Case Series and Quality In Prognosis Studies tools. Synthesis without meta-analysis was conducted. **RESULTS** Sixteen studies (PPCMS+ *n*=252, PPCMS-*n*=590) met criteria for inclusion. Children who experience PPCMS were found to have pronounced, long-term neurocognitive impairments with severely affected processing speed, psychomotor and executive function, and poorer neurocognitive outcomes generally compared to children without PPCMS. Current literature is limited by small samples, lack of diagnostic clarity or routine prospective screening of PPCMS, and limited investigation of factors that may moderate neurocognitive outcomes. **INTERPRETATION** Children with PPCMS have increased vulnerability to neurocognitive impairments which persist beyond the recovery of initial PPCMS symptoms in the post-operative phase. Dedicated research is needed to further our understanding of PPCMS and associated neurocognitive outcomes to inform clinical care.

**What this paper adds:**

- Children who experience PPCMS after surgery experience significant long-term neurocognitive impairment, with most consistent moderate-severe impairments in processing speed, psychomotor function, and executive function.
- Children who experience PPCMS have poorer neurocognitive outcomes generally than children treated for posterior fossa tumour without PPCMS, however future research is needed with larger matched samples of children with and without PPCMS.
- Prospective screening for PPCMS using formal diagnostic criteria in research and clinical practice is recommended, and cognitive development should be monitored in the long-term when PPCMS is identified.
- Future research is needed to understand the role of potential moderating influences on neurocognitive outcomes, such as duration and severity of mutism, age at surgery, and adjuvant oncology treatments.

---

Posterior fossa tumours account for approximately 60% of childhood intracranial tumours. Treatment strategies are predominantly influenced by tumour characteristics, risk classification, and the child’s age, and may include a combination of surgery, chemotherapy, and radiotherapy. Malignant tumours such as medulloblastoma, which account for 50% of posterior fossa tumours, have poorest survival rates necessitating intensive treatment protocols including maximal tumour resection, craniospinal irradiation, and chemotherapy (1). In recent decades, increased survivorship has led to growing recognition of the significant risk of the tumour and associated treatments to long-term cognitive development in childhood survivors, posing a challenge for quality of survival. Post-operative Paediatric Cerebellar Mutism Syndrome (PPCMS), also known as ‘Posterior Fossa Syndrome’ (PFS), is a complication of posterior fossa tumour surgery that presents in approximately 30% of paediatric cases (2–4). PPCMS is characterised by transient mutism, ataxia, hypotonia and emotional lability (5–8). PPCMS symptoms typically onset 1-2 days post-surgery with unpredictable variability in severity and duration, from days to months (9). While the acute cardinal symptoms of PPCMS appear transient, there is increasing evidence that PPCMS may be associated with poorer neurocognitive outcomes in the long-term (10–13), however this remains poorly characterised.

Risk factors for PPCMS have included younger age at tumour diagnosis, medulloblastoma classification, brainstem invasion, and larger tumours that occupy a greater extent of the cerebellar midline (2, 7, 14). The pathophysiology of PPCMS is still incompletely understood. It is hypothesised that the principal cause of PPCMS is damage to the cerebellar outflow pathway which, in the posterior fossa, is just adjacent to a large fourth ventricular tumour and ascends from the deep cerebellar nuclei through the superior cerebellar peduncles and brainstem, via the red nucleus and thalamus to the cortex (15–18). Disruption to this pathway can lead to cerebellocerebral diaschisis and hypoperfusion of the frontal lobe, particularly the supplementary motor areas and limbic system (19–23) resulting in transient motor, speech, and affective symptoms (24). It is suggested that chronic hypoperfusion, reduced oxygen consumption, and hypometabolism of the frontal cortex leads to widespread impact on cognitive function (21, 22, 24).

Despite increasing evidence for persistent neurocognitive impairments associated with PPCMS, it remains unclear if there is a distinct neurocognitive profile of impairment associated with the condition, and if there are moderating clinical and demographic risk factors that could inform risk stratification for neurocognitive outcomes. A systematic review of the neurocognitive outcomes associated with PPCMS is timely as the features and symptoms of PPCMS are now being placed under one definition and label (25) with an increasing need to inform prognostication of long-term outcomes. The current study aimed to systematically review the literature to determine:

1. Evidence for a distinct neurocognitive profile at baseline and longitudinally for children who present with PPCMS.
2. Differences in neurocognitive outcomes between children treated for posterior fossa tumour who present with PPCMS (PPCMS+) compared to those without (PPCMS-).
3. Identification of clinical or demographic factors that moderate neurocognitive outcomes in PPCMS, including:

a. Duration and severity of PPCMS symptoms, which have been associated with other neurological and neuropathological outcomes (18, 26).
b. Age at surgery, tumour classification, adjuvant treatment, and time since diagnosis and surgery, which are commonly associated with neurocognitive outcomes (27–29).

## 1.0. METHOD

A prognostic systematic review was conducted according to the Preferred Reporting Items for Systematic Reviews and Meta-Analysis (PRISMA) statement (30) and protocols (PRISMA-P) (31) and pre-registered on PROSPERO (CRD42024555798).

### 1.1. Eligibility Criteria

Eligibility criteria were informed using the Patients/Population, Intervention/Exposure, Comparator/Control, Outcome (PICO) tool. To be included for review, the study population (P) needed to include children (aged ≤ 18 years) treated with surgical intervention (I) for a posterior fossa tumour with reported standardised neurocognitive assessment data (O). Studies were selected if they compared (C) children who presented with PPCMS (PPCMS+) to children who did not experience PPCMS (PPCMS-) post-operatively, or where neurocognitive performance of a PPCMS+ group was compared to published normative test data.

Cohort studies and case series were included for review if the case series included a minimum of four patients (32). Data for age at neuropsychological assessment in one study (33), and data for neuropsychological tests used in another (34) study could not be retrieved initially but were provided by authors on request. Eligibility criteria are listed in Table 1. The only deviation from pre-registered protocol was to include non-English Language papers for screening to reduce potential bias.

**Table 1.**
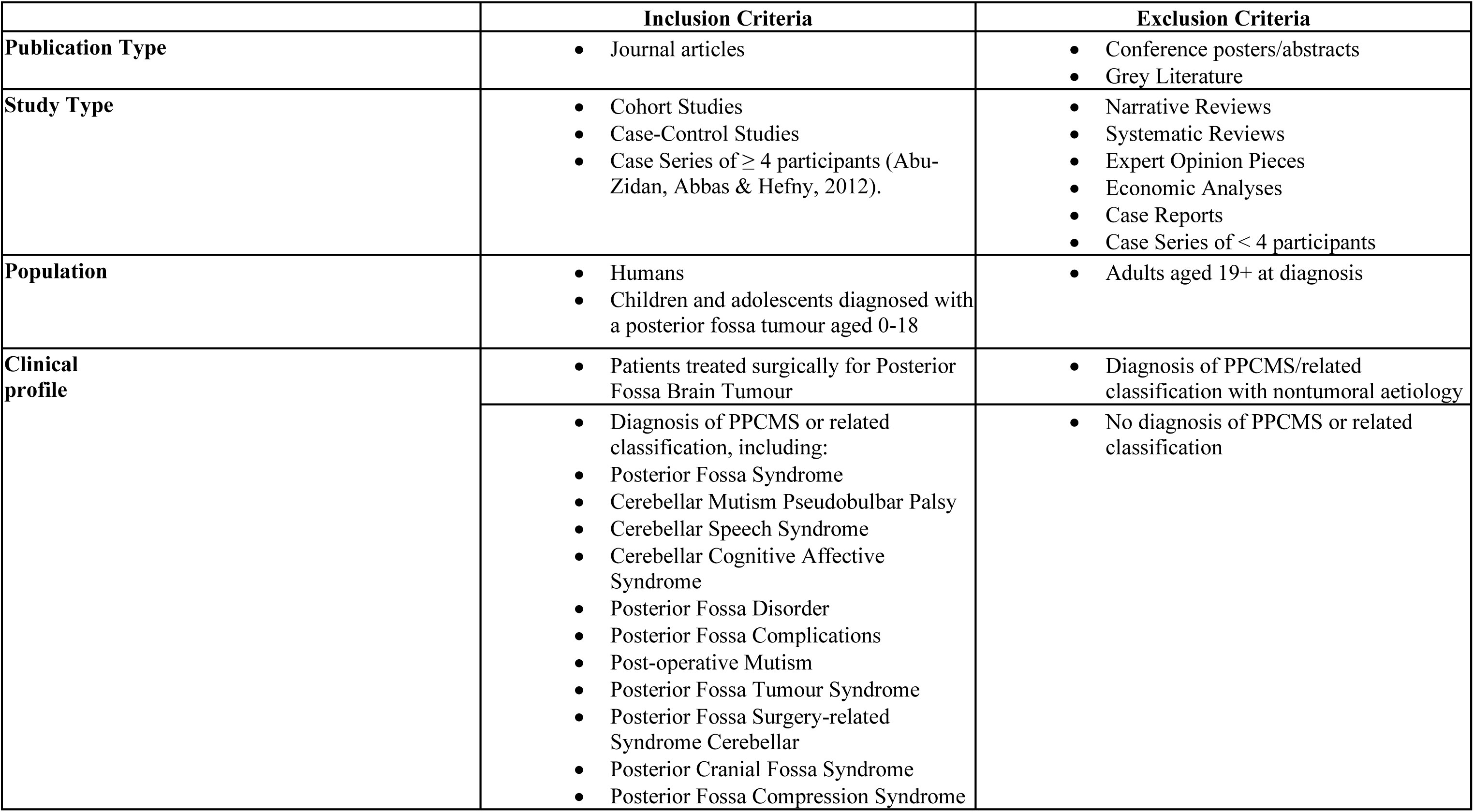
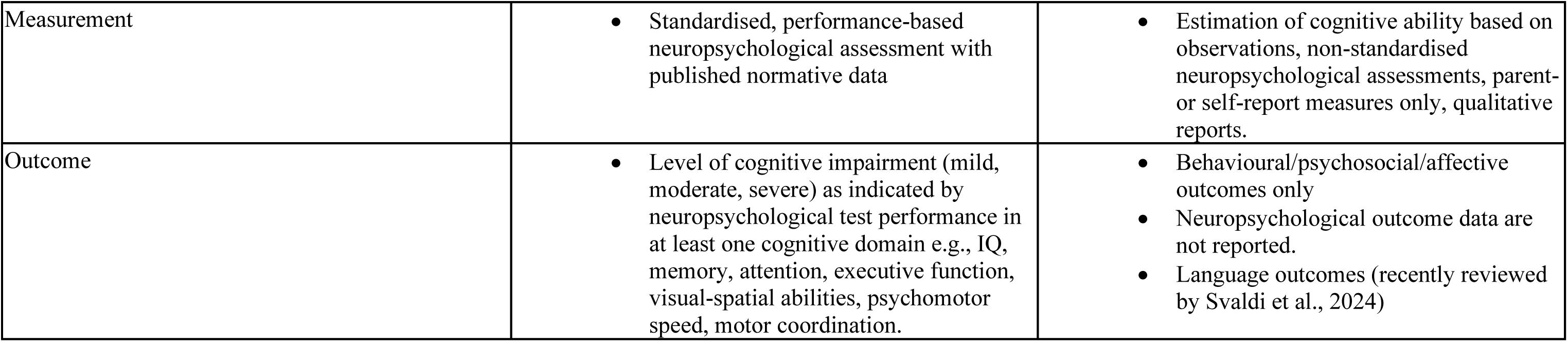
Inclusion and Exclusion criteria for systematic review.

### 1.2. Search Strategy

Embase, PsycInfo, and Medline electronic databases were searched from inception to December 2024. Medical Subject Headings (MeSH) were generated by the National Library of Medicine for each key concept. Key terms were generated from thesauruses and by searching ‘key concepts’ of relevant papers. MeSH terms were connected with key terms for the concepts ‘paediatric’ and ‘cognition’ with ‘OR’. No appropriate MeSH terms were generated for PPCMS as has been the case in previous systematic reviews of PPCMS (2). The final three search terms were searched using ‘AND’. Search terms are listed in Table 2.

**Table 2.** MeSH terms and key terms used in the search.

| Concept | MeSH term | Key term |
| --- | --- | --- |
| Post-operative<br>Cerebellar Mutism<br>Syndrome | Not Applicable | (Posterior Fossa Syndrome or PFS or Cerebellar Mutism or CMS or Pseudobulbar Palsy or Cerebellar Speech Syndrome or Cerebellar Cognitive Affective Syndrome or CCAS or Posterior Fossa Disorder or Posterior Fossa Complications or Posterior Fossa Tumour Syndrome or Posterior Fossa Surgery-related Syndrome or Cerebellar Cognitive Affective Disorder or Posterior Cranial Fossa Syndrome or PPCMS or Post-operative Mutism or Posterior Fossa Compression Syndrome) |
| Paediatric | Pediatric or Pediatrics | (Paed* or Child* or Pediatric* or Adolescen*) |
| Cognition | Cognition or Cognition Disorders or Cognition Assessment or Neuropsychological Tests | (Neurocog* or Neuropsych* or Cogniti* or Intell* or Memory or Executive* or Attention or Processing Speed or Visual Spatial or Visual Motor or Psychomotor) |

### 1.3. Study Selection

Results were exported to Endnote 20 where duplicates were removed. Titles and abstracts of the remaining articles followed by the full texts were then screened by the first author (BMH), with 20% screening by a second reviewer (AAA) according to eligibility criteria (Table 1). Any discrepancies were resolved by a 3^rd^ reviewer (CPM).

### 1.4. Data Extraction

A template for data extraction was created a priori based on pre-registered criteria and piloted with an initial paper. All data were first extracted by the first author (BMH) and validated by a second reviewer (CPM). Data were extracted for 1) study characteristics (e.g., authors, year of publication, study type, sample sizes), 2) neurocognitive domains, assessment measures, and outcomes, and 3) details of analysis for potential clinical and demographic moderators or co-variates.

### 1.5. Quality Assessment

All included studies were assessed for methodological quality by two independent reviewers (BMH & AAA). The quality of included case series was assessed using the Institute of Health Economics (IHE) Quality Appraisal Checklist for Case Series Studies (35) and cohort studies were assessed using the Quality In Prognosis Studies (QUIPS) tool (36). 20% of quality appraisals were checked, and any discrepancies resolved by a third reviewer (TM).

### 1.6. Data Synthesis

Due to significant variability in the cognitive assessment measures used and associated metric reported (e.g., standard score, T-score, percentile) between studies, a meta-analysis or one sample *t*-test was not appropriate, and synthesis without meta-analysis (SWiM, 37) was conducted. The data were synthesised using pre-registered categories:

#### Study and Population Characteristics

Year of publication and number of PPCMS+ and PPCMS-posterior fossa tumour patients in each study.

#### Neurocognitive Assessment

The neurocognitive tests used, and cognitive domains assessed (see Table 3). Language outcomes were not included as they have recently been systematically reviewed (38).

#### Neurocognitive Outcomes

1. Standardised cognitive test scores. Neurocognitive outcomes were grouped conventionally according to standard deviation (SD) distance from the normative mean, with impairment as either mild (1-1.5 SD below the mean), moderate (1.5-2 SD below the mean) or severe (>2 SD below the mean). The average level of impairment reported by domain was used to synthesise data from cohort studies. The average level of impairment was used for case series where an average was reported, or alternatively by individual cases. Cognitive data were grouped according to time since surgery. Baseline assessments were classified as those conducted pre-operatively or within 6 months post-operatively, and long-term outcome assessments were classified as at least 12 months post-operatively.
2. Report of statistical comparison of neurocognitive outcomes between PPCMS- and PPCMS+ patients.

#### Clinical and Demographic Factors

Report of statistical associations between neurocognitive outcomes for PPCMS and duration and severity of mutism, tumour and adjuvant treatment type, age at diagnosis and surgery, and time since diagnosis and surgery.

**Table 3.** Description of neurocognitive domains reviewed.

| Neurocognitive domain | Explanation |
| --- | --- |
| General intellect | An individual's general cognitive ability (e.g., Full Scale IQ) |
| Verbal Intellect | General knowledge and reasoning using verbal information and concepts. |
| Non-Verbal Intellect | General knowledge and reasoning using non-verbal information and concepts. |
| Short-term and Working Memory | Short-term memory is the ability to hold verbal and visual information in short-term stores (~30 seconds).<br>Working memory is the ability to manipulate information in short-term stores to perform a computation. |
| Processing Speed | The speed at which an individual can receive, process, and respond to information. |
| Long-term memory | Encoding, storage, and retrieval of visual or verbal information after a delay (>30 seconds). |
| Executive Function | A set of related cognitive skills for controlling and monitoring behaviour to achieve a goal. Including planning and organising information, problem-solving, decision-making, monitoring behaviour, response inhibition, and switching flexibly between information. |
| Attention | Selective attention: Prioritising focus to stimuli over irrelevant or competing information.<br>Sustained attention: Maintaining focus over prolonged, repetitive, or monotonous activity.<br>Focused attention: The basic ability to orient to a single stimulus for a short period. |
| Psychomotor Function | Accuracy and speed of a motor response proceeding cognitive activity e.g., 'hand-eye' co-ordination after perceiving an object. |
| Visual-Spatial Abilities | The capacity to perceive, evaluate, and mentally manipulate visual details and spatial relationships. |

## 2.0. RESULTS

### 2.1. Study Selection and Quality Appraisal

The initial search yielded 1922 articles with 1,503 remaining following duplicate removal. After screening abstracts and titles, 132 articles remained (97.27% inter-rater agreement). Two non-English language papers were included for full-text screening and translated using a machine translation engine (Google Translate) but did not remain eligible against inclusion and exclusion criteria (one case study and one did not conduct neuropsychological assessment). Sixteen studies remained eligible after full-text screening against inclusion and exclusion criteria (100% inter-rater agreement). A Flow Diagram of the search process is presented in Figure 1.

**Figure 1:**
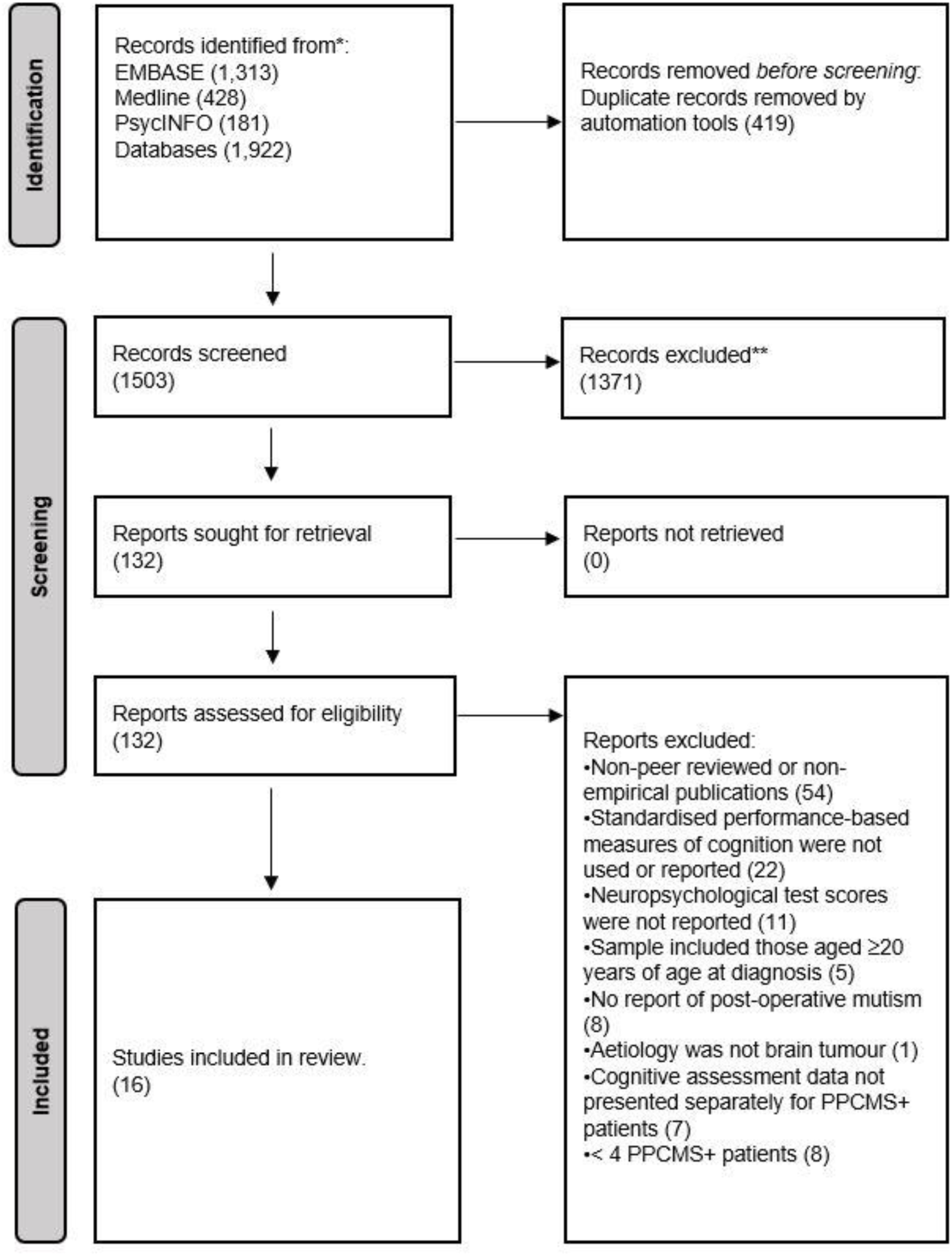
PRISMA Flow Diagram.

The methodological quality of prognostic cohort studies is summarised in Appendix S1. Risk of bias was assessed to be mostly moderate, with no studies assessed to have high risk of bias (100% inter-rater agreement). Methodological quality was limited by study attrition due to severity of PPCMS symptoms restricting cognitive assessment (39) or no report of the characteristics of those lost to follow-up (40, 41). Whilst most studies used high quality measurement or classification of tumour characteristics and medical treatments, the methodological quality of cohort studies was limited by lack of clear and consistent measurement and diagnosis of PPCMS. The cognitive assessment of PPCMS was predominantly a secondary variable of interest (in 69% of studies) rather than central to the study aims. Similarly, the methodological quality of case series was limited by exclusion of those with severe PPCMS symptoms (42) and incomplete neuropsychological follow-up.

Notably three cohort studies had planned to conduct baseline assessments as part of a pre-determined trial protocol but were either unable to or excluded patients with PPCMS at baseline due to the severity of symptoms prohibiting assessment (39, 43, 44). Only one study assessed cognition pre- and post-operatively, however the authors were not able to provide separate standardised neurocognitive data for their PPCMS+ and PPCMS-groups at pre-operative baseline (42).

### 2.2. Study Characteristics

Characteristics of included cohort studies and case series are summarised in Table 4 and 5 respectively. The *n* for each study reflects the number of patients with neurocognitive data reported, which is the same as the study total sample size except for two studies (34, 39). Eleven studies referred to PPCMS as ‘Cerebellar Mutism’ (33, 34, 41–49) five referred to PPCMS as ‘Posterior Fossa Syndrome (39, 40, 50–52). No other terms for PPCMS were used.

**Table 4.**
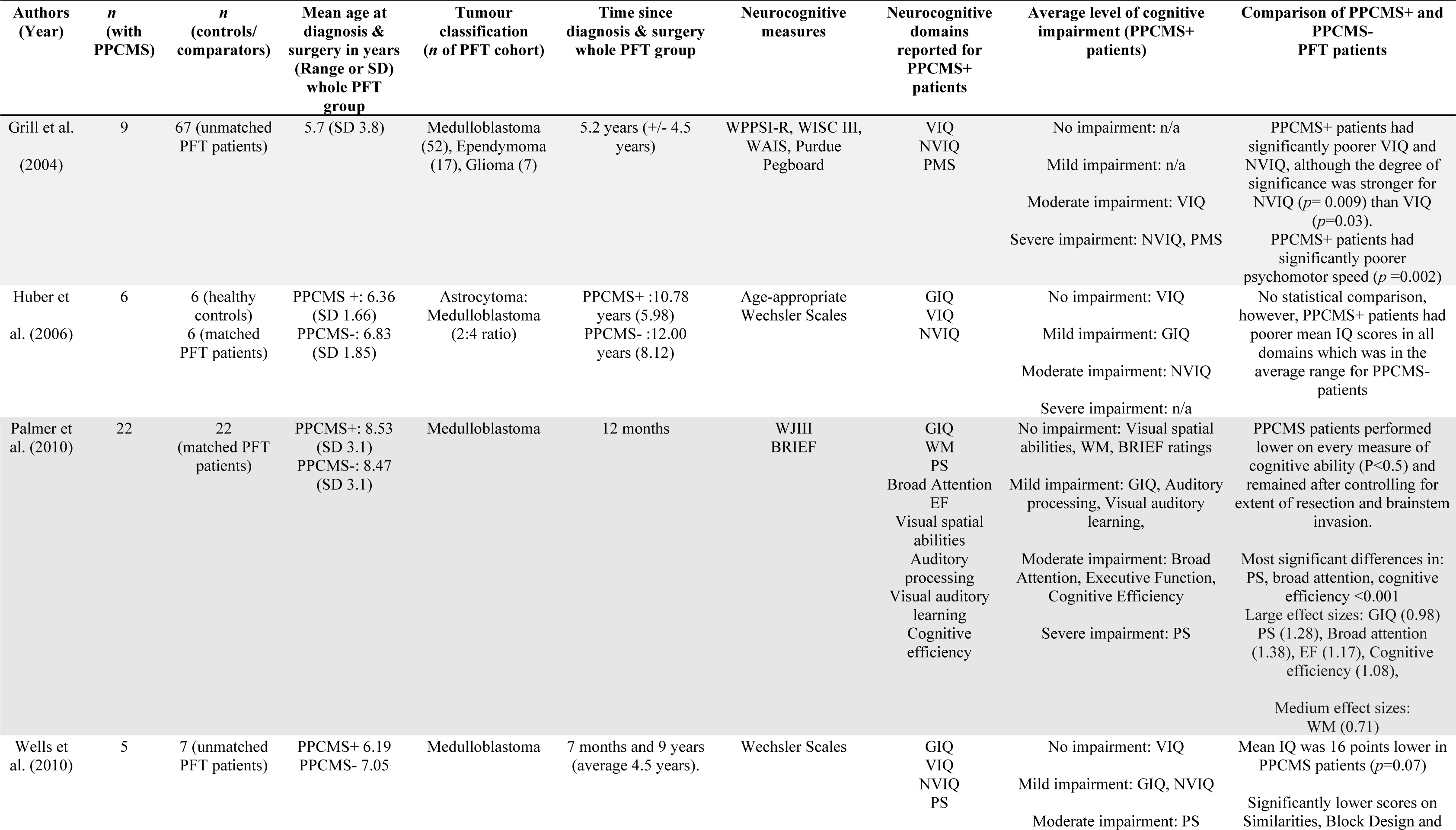
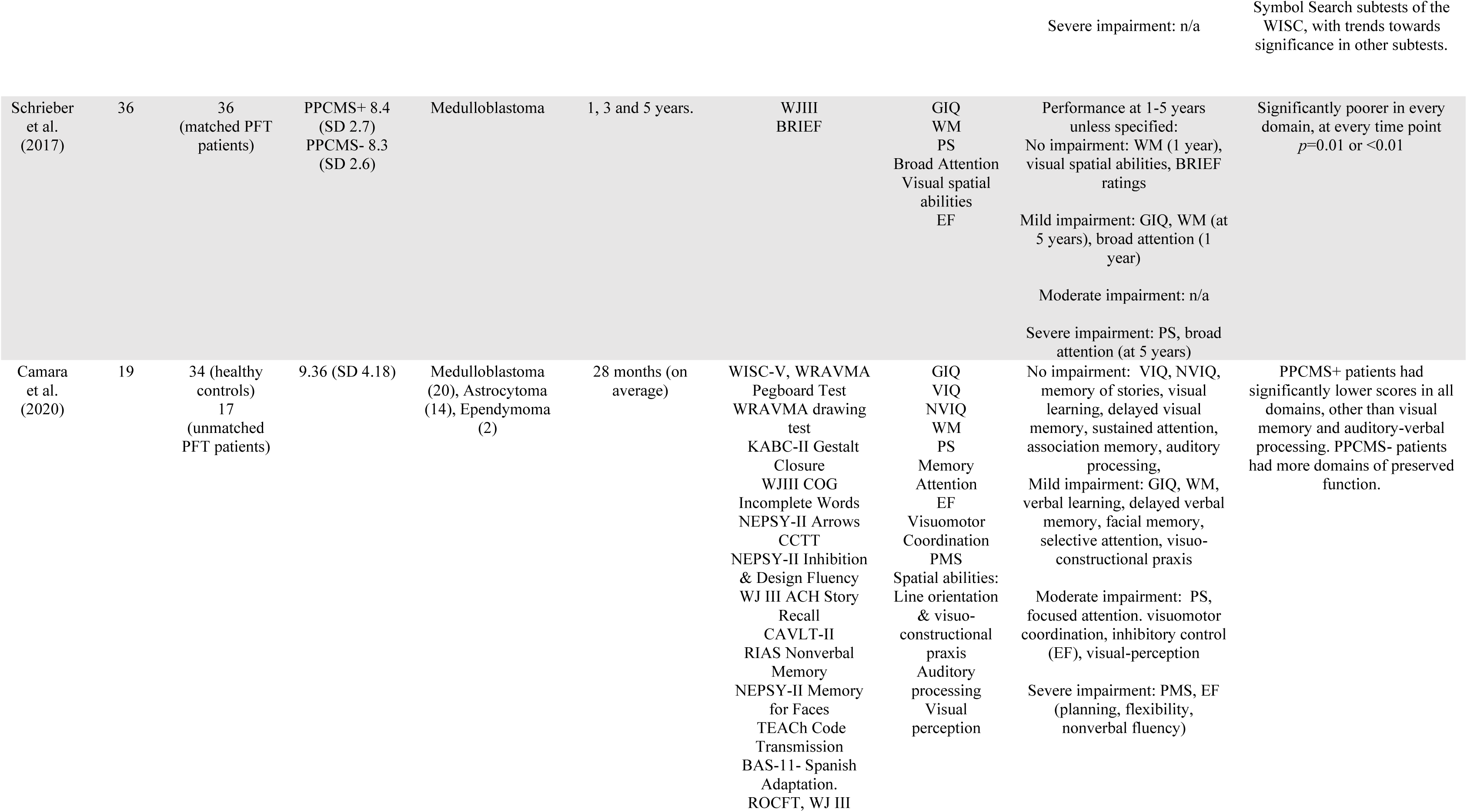
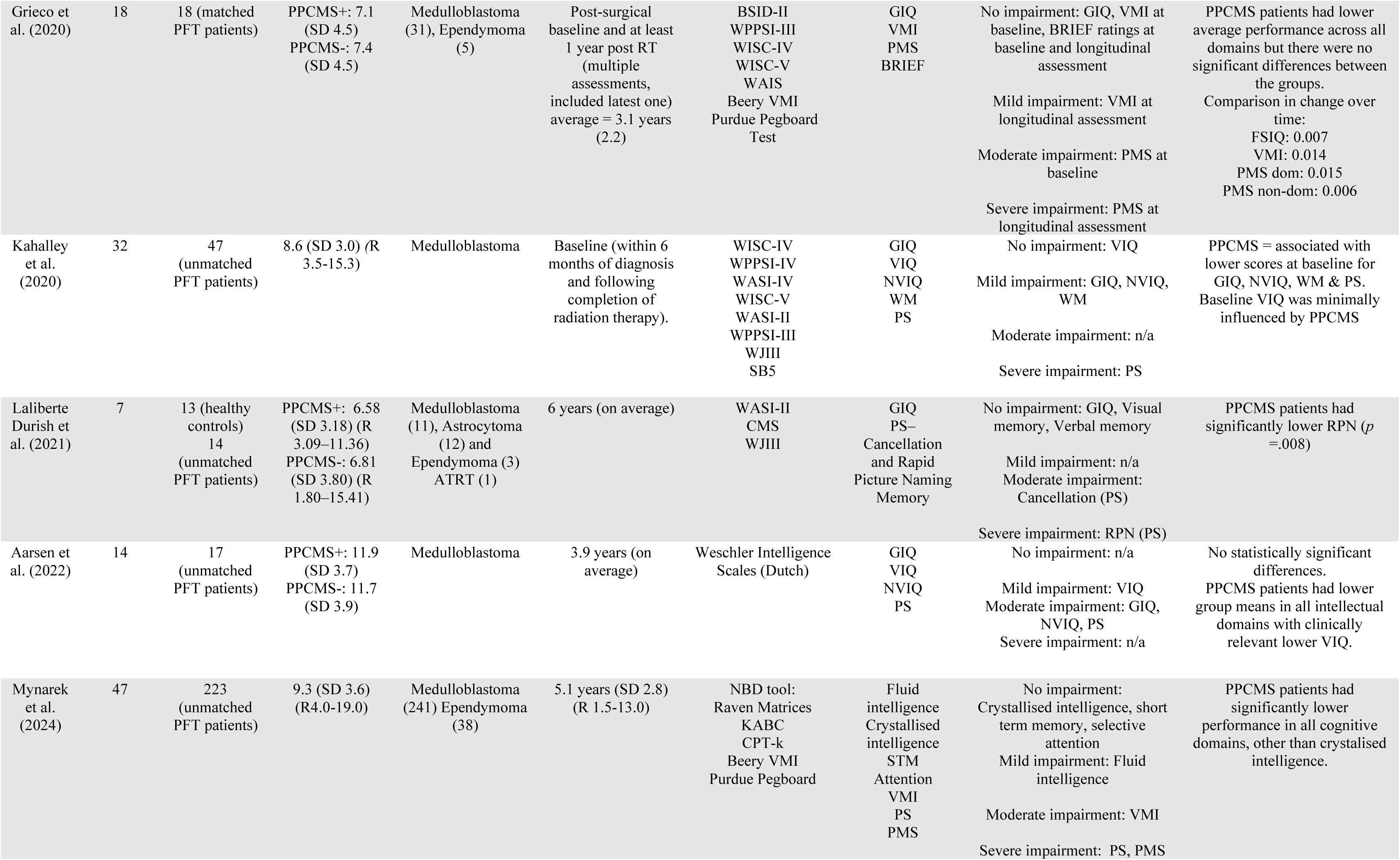
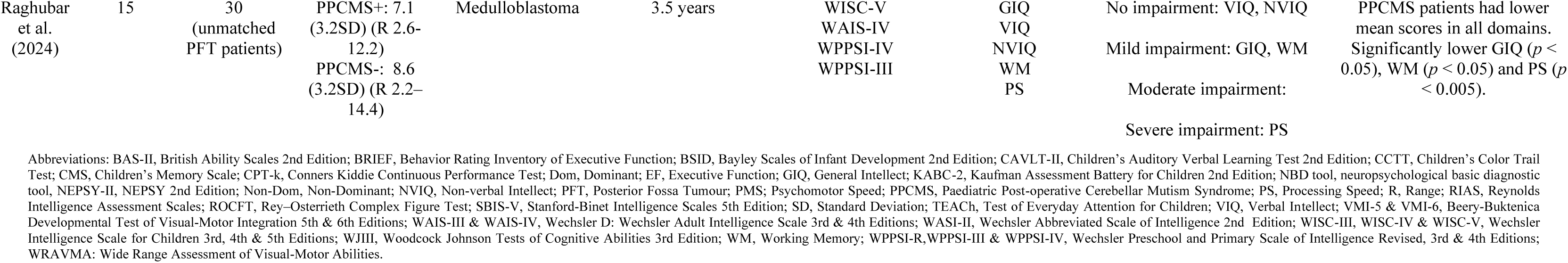
Characteristics of cohort studies reviewed.

**Table 5.**
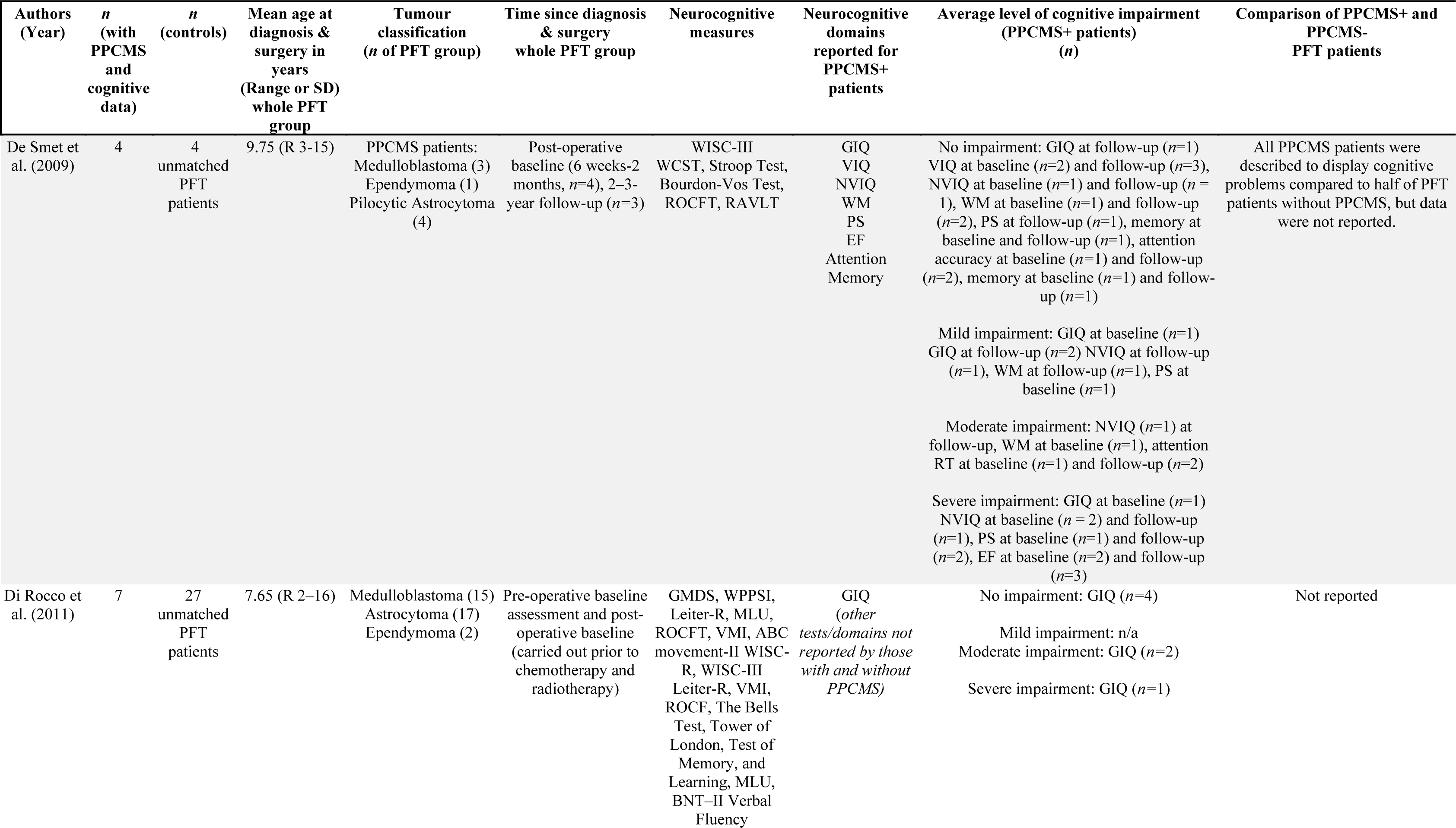
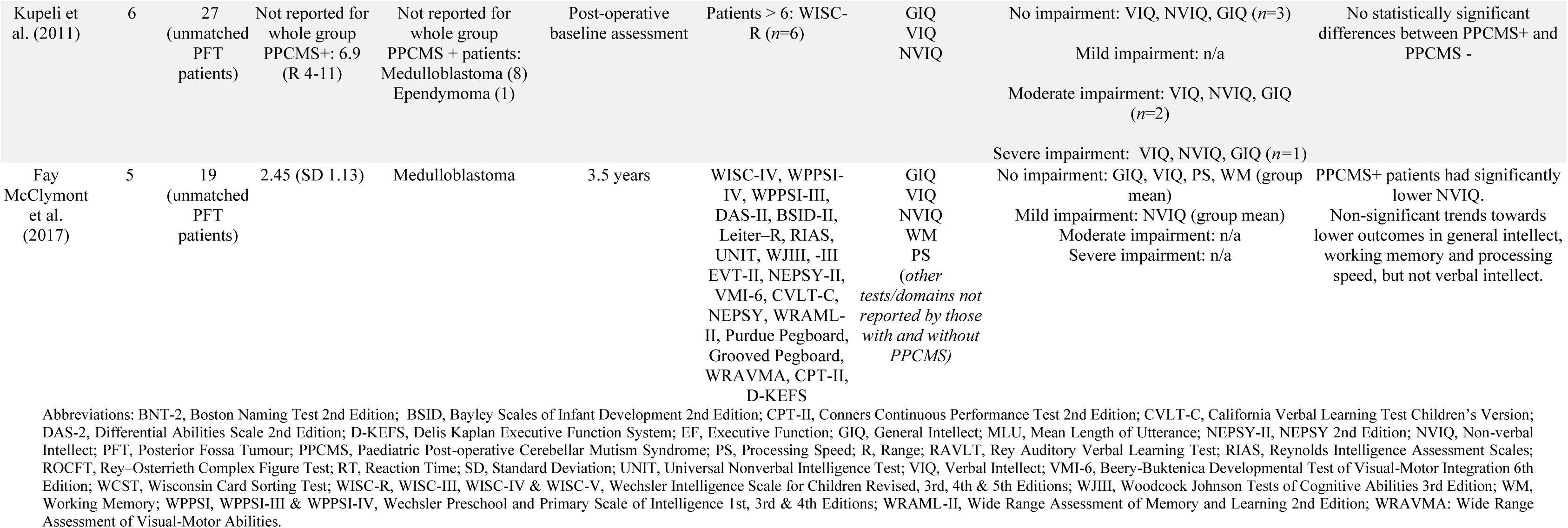
Characteristics of case series reviewed.

#### 2.2.1 Cohort Studies

Twelve cohort studies published between 2004 and 2024 were included for review, with performance-based neurocognitive data for a total of 230 children with PPCMS and 513 children without PPCMS (33, 34, 39–41, 43–49). Two cohort studies reported neurocognitive outcomes at baseline (40, 44, *n*=50). Eleven cohort studies reported long-term neurocognitive outcomes from 1-13 years post-tumour diagnosis (33, 34, 39, 41, 43-48, 49, *n*=198). Cohort studies reported on long-term neurocognitive outcomes in the following domains (number of studies): general intellectual function (=9), processing speed (=8), verbal intellect (=7), non-verbal intellect (=7), working memory (=4), attention (=4), psychomotor function (=4), long-term memory (=3), visual spatial abilities (=3), and executive function (=2).

#### 2.2.2. Case Series

Four case series published between 2009 and 2017 were included for review with a total of 22 children with PPCMS and 77 without PPCMS (42, 50–52). Three case series reported neurocognitive outcomes at baseline (42, 50, 51) and two reported long-term neurocognitive outcomes 2-3.5 years post-tumour diagnosis (50, 52). Three case series reported intellectual outcomes only (42, 51, 52, *n*=18) and one case series (*n*=4) reported test scores for executive and intellectual functions (50).

### 2.3. Neurocognitive outcomes

#### 2.3.1. Baseline

Two cohort studies (*n*=50) reported cognitive data for PPCMS at baseline. One study reported age-appropriate verbal intellect, mild impairments in general intellect, non-verbal intellect, and working memory, and severe impairments in processing speed (40, *n*=32). The other reported age-appropriate general intellect (individual indices of intellect were not reported), age-appropriate visual-motor integration skills, and moderate impairments in psychomotor speed (44, *n*=18).

Three case series (42, 50, 51, *n*=17) also reported similar mixed findings for post-operative baseline intellectual abilities. One case series (*n*=4) reported severe impairment in executive functioning at baseline (50).

#### 2.3.2 Long-term neurocognitive outcomes for PPCMS+ patients

**Figure 2:**
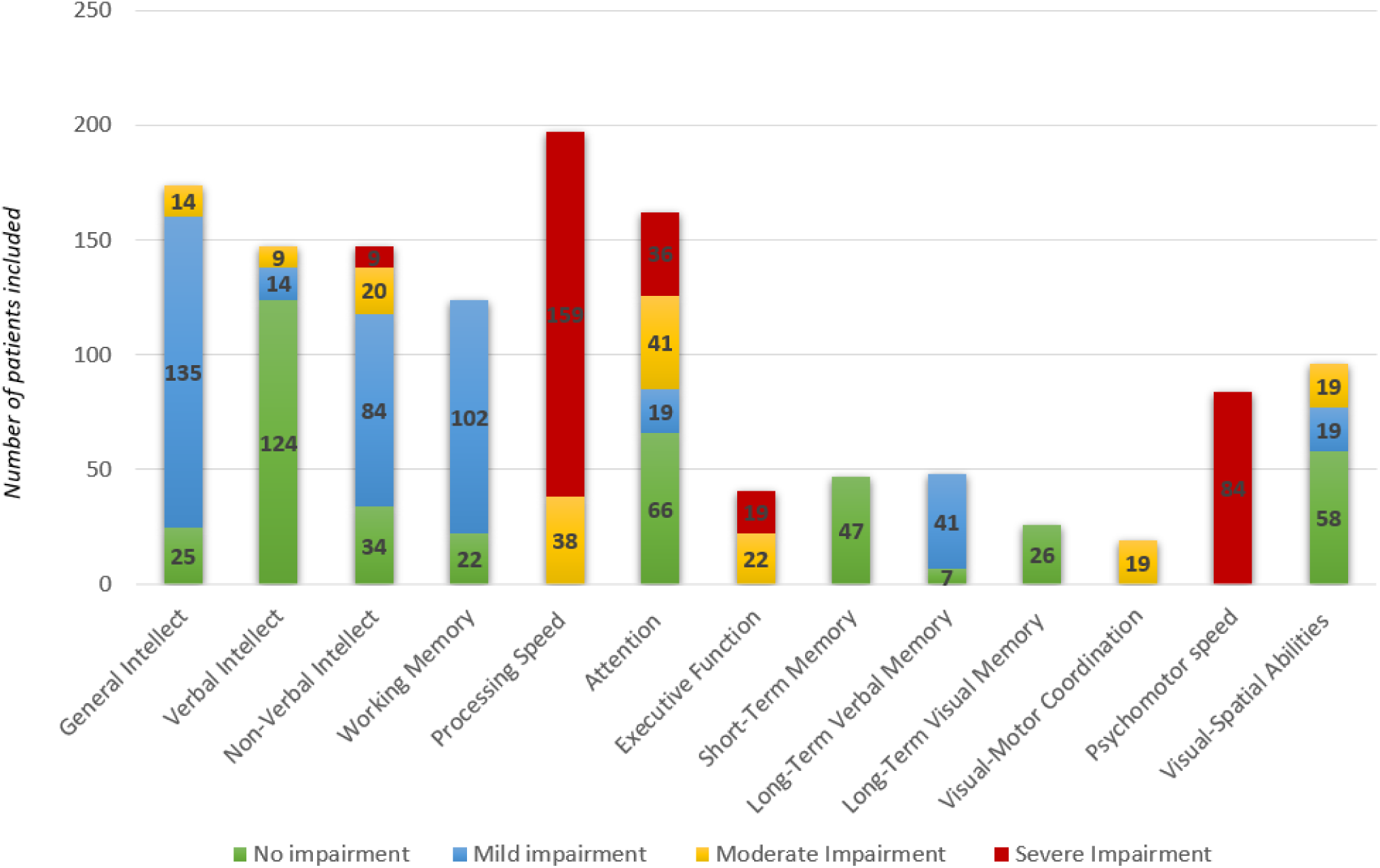
Level of neurocognitive impairment per domain reported across cohort studies. The n reflects the number of patients included in each cohort study that reported a mean score in the severe, moderate, mild, or no impairment range for the PPCMS+ group. No individual patient data is reported. Only performance-based cognitive data is reflected in this figure. ‘Attention’ reflects data for sustained selective, focused, and broad attention.

##### General Intellect

Nine cohort studies (*n*=142) reported on general intellectual function, of which only two reported no impairment (44, 45, *n*=25), six reported mild impairments (34, 39, 43, 46, 47, 49, *n*=103), and one reported moderate impairment (41, *n*=14). Long-term outcomes in general intellect were reported on by both case series (*n*=9). One case series reported no impairments (52, *n*=5) and the other reported mixed findings ranging from no to mild impairment (50, *n*=4).

##### Verbal Intellect

Seven cohort studies (*n*=115) investigated verbal intellect; five of which assessed verbal intellect to be age-appropriate (33, 34, 46, 47, 49, *n*=92). Of the remaining two studies, one reported mild impairments (41, *n*=14) and one reported moderate impairments (48, *n*=9). Long-term verbal intellect was reported to be age-appropriate by case series (50, 52, *n*=9).

##### Non-Verbal Intellect

Non-verbal intellect was investigated by seven cohort studies (*n*=115) with mixed findings. One study reported severe impairments (48, *n*=9), two reported moderate impairments (41, 49, *n*=20), two reported mild impairments (33, 34, *n*=52), and two reported no impairments (46, 47, *n*=34). Mixed findings were also reflected within case series. One case series reported mild impairments (52, *n*=5), and the other reported mixed findings across the severity spectrum (50, *n*=4).

##### Short-Term and Working Memory

Working Memory was investigated by four cohort studies (*n*=92). Three reported mild impairments (39, 46, 47, *n*=70), and one study reported no impairment (43, *n*=22). One case series investigated working memory and reported no impairments (52, *n*=5). One study reported no impairments in short-term memory (33, *n*=47).

##### Processing Speed

Eight cohort studies assessed processing speed (*n*=165). Five (*n*=127) reported severe impairments (33, 39, 43, 45, 47) and three (*n*=38) reported moderate impairments (34, 41, 46). One case series investigated processing speed and reported no impairments but only reported data for 5 patients (52).

##### Long-Term Memory

Memory outcomes were reported by three cohort studies (*n*=48), of which one reported no impairments in visual memory and mild impairments in verbal memory (46, *n*=19), another reported no impairments in visual and verbal memory (45, *n*=7) and another reported mild impairments in visual-auditory memory (43, *n*=22). Memory outcomes were reported for one patient in one case series and noted to be average (50).

##### Psychomotor Function

Psychomotor Speed was investigated by four cohort studies, which all reported severe impairments (33, 44, 46, 48, *n*=93). One cohort study investigated visuomotor coordination and reported moderate impairments (46, *n*=19). Psychomotor functions were not investigated by case series.

##### Executive Function

Executive function outcomes on performance-based neurocognitive measures were investigated by two cohort studies (*n*=41). One reported moderate impairments (43, *n*=22) and the other reported severe impairments in planning, flexibility, and fluency, although it is noted that all tests reported on involved a motor response (46, *n*=19). One case series reported severe impairments in verbal executive function (50, *n*=4). Parent-report of executive functions on standardised questionnaire measures were also reported by three cohort studies to be age-appropriate in notable contrast to performance-based measures (39, 43, 44, *n*=76).

##### Attention

Four cohort studies investigated attention (*n*=124). One study reported no impairment in selective attention (33, *n*=47). Another study reported no impairment in sustained attention, mild impairment for selective attention, and moderate impairment for focused attention which were measured using processing speed tasks (46, *n*=19). One study (43, *n*=22) reported moderate impairments for a multidomain index of attention 1-year post-surgery. Another study (39, *n*=36) reported mild impairment for a multidomain index of attention 1-year post-surgery and severe impairment 5 years post-surgery. Attention was reported by one case series (50) for two patients indicating moderately impaired reaction times but average accuracy.

##### Visual-Spatial/Perceptual Abilities

Visual-spatial cognition was reported by three cohort studies (*n*=77), with two reporting no impairments for spatial relations tasks (39, 43, *n*=58), and one reporting mild impairments for visuo-constructional praxis, and moderate impairments in visual-spatial and perceptual tasks (46, *n*=19). Visual-spatial abilities were not reported by case series.

##### Other Domains

Two studies reported neurocognitive outcomes in additional cognitive domains, with one reporting mild impairments in auditory processing and moderate impairments in cognitive efficiency (43, *n*=22), and the other reporting no impairments in auditory processing (46, *n*=19).

### 2.4. Comparison between children with and without PPCMS

All 12 cohort studies compared neurocognitive outcomes between children with PPCMS (*n=*230) and those without (*n*=513). Seven reported PPCMS+ patients to have statistically significant lower scores in all cognitive domains, other than in domains where no impairment was found for either group, such as verbal intellect (33, 34, 39, 43, 46, 47). The largest effect sizes for group differences were reported in processing speed, attention, general intellect, and executive function with medium effect sizes in working memory (43). The four studies that did not find significant differences between PPCMS+ and PPCMS-groups reported trends towards lower scores in all cognitive domains for children with PPCMS (40, 41, 44, 49).

Three case series compared neurocognitive outcomes for children with PPCMS (*n*=15) and those without (*n*=50). One reported significantly poorer non-verbal intellect, and non-significant trends towards lower outcomes in general intellect, working memory and processing speed, but not verbal intellect in their PPCMS+ group (52). Another qualitatively described poorer outcomes for children with PPCMS with insufficient sample size for statistical comparison (50). One case series reported no qualitative or statistically significant differences between PPCMS+ (*n*=6) and PPCMS-groups (*n*=27) (51).

### 2.5. Associations with Clinical and Demographic Factors

Associations between neurocognitive outcomes in PPCMS with clinical and demographic risk factors are described for cohort studies and case series together below and summarised in Table 6.

**Table 6.** Associations between clinical and demographic risk factors with neurocognitive outcomes for PPCMS+ patients.

| Factors | Reported for PPCMS+ patients | Summary of characteristics | Statistical analysis with cognition for PPCMS+ patients as a subgroup |
| --- | --- | --- | --- |
| **Mutism Duration** | Yes: 5 studies (*n*=40)<br>No: 11 studies (*n*=215) | ≤ 1 week: (*n*=16)<br>1-4 weeks: (*n*=11)<br>1-2 months: (*n*=8)<br>4-6 months: (*n*=3)<br>> 6 months: (*n*=2)<br>(total *n*: Huber et al., 2006; Aarsen et al., 2022; De Smet et al., 2009; Di Rocco et al., 2011; Kupeli et al., 2011) | Variable analysed: 1 study (Aarsen et al., 2022, *n*=14).<br>No association with cognition<br>Variable not analysed: 15 studies. |
| **Mutism Severity** | Yes: 3 studies (*n*=26)<br>No: 13 studies (*n*=229) | Mild: *n*=4, Moderate: *n*=9, Severe: *n*=8 (Aarsen et al., 2022; Di Rocco et al., 2011)<br>Wells et al. (2010) reported 82% of sample to have moderate/severe PPCMS symptoms | Variable not analysed: 16 studies. |
| **Mean age at diagnosis of posterior fossa tumour and surgery** | Age at diagnosis was reported for PPCMS+ patients separately:<br>Yes: 10 studies (*n*=135)<br>No: 6 studies (*n*=120) | 6 years: 4 studies (*n*=32)<br>7 years: 2 studies (*n*=16)<br>8 years: 4 studies (*n*=87)<br>Total range for all studies: 2.6-15.3 years<br>No report of age: 6 studies (*n*=120) | Variable not analysed: 16 studies. |
| **Average time since treatment** | Yes: 10 studies (*n*=171)<br>No: 6 studies (*n*=84) | Baseline only: 3 studies (*n*=48)<br>1 year: 1 study (*n*=22)<br>2 years: 2 studies (*n*=23)<br>3 years: 2 studies (*n*=29)<br>6 years: 1 study (*n*=7)<br>10 years: 1 study (*n*=6)<br>1, 3 & 5 years: 1 study (*n*=36)<br>Total range for all studies: 1-13 years | Variable analysed: 2 studies.<br>Grieco et al. (2020) (*n*=18) non-significant standard score declines in mean GIQ, VMI & PMS<br>Schrieber et al. (2017) (*n*=36) significant standard score declines in attention. Nonsignificant declines in WM<br>Variable not analysed: 14 studies |
| **Tumour classification** | Yes: 14 studies (*n*=199)<br>No: 2 studies (*n*=56) | Medulloblastoma: *n*=176<br>Astrocytoma: *n*=16<br>Ependymoma: *n*=7 | Variable not analysed: 16 studies. |
| **Adjuvant treatments** | Yes: 9 studies (*n*=162)<br>No: 7 studies (*n*=93) | Surgery + radiotherapy + chemotherapy: *n*=133<br>Surgery+ radiotherapy: *n*=6<br>Surgery only: *n*=23 | Variable analysed: 2 studies.<br>- Kahalley et al. (2020) PPCMS is associated with lower performance in all areas of intellectual function, independent of RT type.<br>- Mynarek et al. (2024) The association between RT type (high-dose CSI versus focal RT) and PS was no longer significant with PPCMS as a co-variate.<br>Variable not analysed: 14 studies. |

Abbreviations: CSI, Craniospinal Irradiation; GIQ, General Intellect; PMS, Psychomotor Speed; PS, Processing Speed; PPCMS, Paediatric Post-operative Cerebellar Mutism Syndrome; RT, radiotherapy; VMI, Visual-motor integration.

#### Duration of Mutism

One cohort study directly investigated the association between duration of PPCMS with neurocognitive outcomes and found no significant association with intellectual outcomes, the only cognitive functions investigated in this study (41, *n*=14). Case series data described patients who experienced more than 1 week of mutism (*n*=5) had more severe neurocognitive outcomes, although this was not statistically analysed (42, 50).

#### Severity of Mutism

Severity of PPCMS was reported by two cohort studies (34, 41) and one case series (42) but no studies statistically analysed the relationship between mutism severity and cognition. One cohort study reported that the exclusion of patients with severe PPCMS at baseline meant the relationship between PPCMS symptoms and cognition could not be investigated (39, *n*=36).

#### Age at diagnosis/surgery

No studies statistically analysed the relationship between age at diagnosis or surgery and neurocognitive outcomes for PPCMS+ patients separately from PPCMS-patients.

#### Time since diagnosis and surgery

One study reported significant declines in standard scores between baseline and, on average, 3-years post-diagnosis in mean general intellect, visual-motor integration, and psychomotor speed for PPCMS+ patients (44, *n*=18). Another study reported significant declines in a multidomain index of attention and nonsignificant declines in working memory between baseline and 5 years post-surgery for the PPCMS+ group (39, *n*=36).

#### Tumour Classification and Adjuvant Treatments

Fourteen studies (*n*=199) reported the specific tumour classification for PPCMS+ patients, with medulloblastoma diagnosis in 88% of cases. It should be noted that seven studies only included patients treated for medulloblastoma, which leads to potential bias of tumour classification, although the prevalence of PPCMS is also significantly higher for medulloblastoma. No studies investigated the relationship between tumour classification on neurocognitive outcomes for PPCMS.

Treatment type was reported separately for PPCMS+ patients by nine studies (*n*=162). Most patients were treated with radiotherapy and chemotherapy after surgery. One study found PPCMS+ patients to perform lower in all areas of intellectual function, independent of radiotherapy treatment modality (craniospinal photon versus proton-beam radiotherapy) (40, *n*=32). Another study reported that radiotherapy treatment type and dose (high-dose craniospinal irradiation versus focal radiotherapy) no longer accounted for variance in processing speed when PPCMS was added as a covariate (33, *n*=47). No other studies statistically analysed the relationship between exposure to adjuvant treatments and neurocognitive outcomes in PPCMS.

#### Additional factors

One study reported larger tumour size, presence of preoperative hydrocephalus and higher mean basal temperature 1–4 days after surgery to be predictors for lower general intellect, verbal intellect, and processing speed for their PPCMS+ patients (41, *n*=31). In a small sample, decline in fine motor speed and coordination for both PPCMS+ and PPCMS-groups was associated with subtotal resection (44, *n*=36).

Two studies reported statistically significant differences in clinical variables between PPCMS+ and PPCMS-groups, however, these factors were not analysed as co-variates and may influence group differences otherwise attributed to PPCMS. This includes a higher number of surgeries (39, *n*=72) and greater frequency of hydrocephalus requiring a shunt (47, *n*=45) in the PPCMS+ groups.

## 3.0 DISCUSSION

To our knowledge, this is the first systematic review of neurocognitive outcomes in PPCMS. The search identified sixteen empirical papers for inclusion and revealed a near doubling of relevant papers in the past five years, reflecting an increased growth in acknowledgement of longer-term consequences of PPCMS justifying systematic review.

### Neurocognitive profile of PPCMS

The review aimed to establish evidence for a distinct neurocognitive profile for children who present with PPCMS. Across studies, PPCMS was associated with significant long-term cognitive impairment in multiple domains. Processing speed, psychomotor function, and executive function were the domains with most consistent moderate-severe impairment and a markedly high rate of mean severe impairment. These domains are particularly reliant on integrity of the frontal lobe and cerebrocerebellar pathways (53–55) which is supportive of current hypotheses of PPCMS pathophysiology (17, 21). The consistent findings of moderate-severe long-term psychomotor deficits across studies support the view of persistent impairment in psychomotor function even after some resolution of acute ataxia (33). Assessment of processing speed usually requires a visual-motor response, and it is plausible that psychomotor deficits contribute to the prevalence of impairment in processing speed, however this is unlikely to be the sole contributor (see 56). Processing speed is an integrative process involving cognitive speed and proficiency, executive control, and the efficient integration of sensory, motor, and cognitive processes (57, 58), therefore drawing on multiple domains and underlying neural networks that appear vulnerable to impairment after PPCMS. Consistent severe impairment in processing speed potentially reflects the cumulative effect of multidomain deficits. However, it will be valuable for future research to include motor-free or motor-reduced assessment of processing speed and executive function (56) to better characterise this grouping of marked deficits. Similarly, it will be valuable for future research to explore the discrepancy found between caregiver report and performance-based measurement of executive function, particularly where there are limits to direct assessment of executive function in pre-adolescence as skills emerge. The findings emphasise the need for long-term neurocognitive monitoring after PPCMS to assess vulnerable higher-order cognitive processes as they undergo rapid development in adolescence.

Most preserved and average range function was found for verbal intellect and long-term memory. Verbal intellect is often relatively preserved for children treated for posterior fossa tumours but only represents semantic aspects of language, whilst other domains of language skills are found to be more frequently impaired after PPCMS (38). Long-term memory was insufficiently studied to confidently conclude absence of impairment and should be a focus of further study given suggested limbic involvement in PPCMS.

The greatest variability of findings was for non-verbal intellect and attention, with mixed findings from severe difficulties to preserved function. Differences within these domains may be due to variability in the underlying component constructs and neuropsychological tests used to assess these skills over time and between studies. For example, non-verbal fluid reasoning and visual-spatial cognition were previously combined as a single index of non-verbal intellect on earlier Wechsler scales of intelligence but have subsequently been separated into distinct indices to provide greater interpretive clarity of clearly distinct domains. As fluid reasoning tasks usually demand greater executive function than tasks of visual-spatial manipulation (59, 60), mixed findings for non-verbal intellect may be the result of grouping these disparate tasks under a single domain. This is consistent with previous interpretation of preserved visual-spatial ability in PPCMS when tasks administered were multiple-choice with reduced demands on planning and organisation (39). Similarly, attentional skills are multidimensional (e.g., sustained versus selective attention) and mediated by multiple neural networks across childhood (61, 62). Further research is needed to determine which attentional skills are impacted after PPCMS, although there is clear evidence of vulnerability to impaired attention that increases over development after PPCMS (39).

There were insufficient data to reliably characterise the baseline neurocognitive profile of PPCMS with certainty. This is a common challenge in paediatric neuro-oncology due to rapid treatment required at diagnosis, and post-operative symptoms prohibiting valid baseline neurocognitive assessment. It is recommended that standardised caregiver report of pre-operative development is used in research and clinical practice where performance-based testing at baseline is not possible, rather than excluding children with PPCMS from study. This approach can provide valuable information about baseline neuropsychological risk factors to aid both risk stratification and prognostication of long-term outcomes when measures can be repeated.

### Comparison between children with and without PPCMS

The second aim of the review was to determine any difference in neurocognitive outcomes between children who present with and without PPCMS. A methodological strength of the included studies was the consistent comparison of neurocognitive outcomes between PPCMS+ and PPCMS-groups in all cohort studies and three case series. Children with PPCMS consistently displayed poorer neurocognitive outcomes in all impaired cognitive domains, and particularly in processing speed, attention, executive function, working memory and non-verbal intellect. These findings support the hypothesis that children who experience PPCMS have either a broader or additional neuropathology, potentially resultant of cerebellocerebral diaschisis (17, 21, 22, 24).

### Associated clinical and demographic risk factors

The final aim of the review was to identify any moderating influence of clinical and demographic factors on neurocognitive outcomes for children with PPCMS. Duration and severity of mutism, age at diagnosis and surgery, time since surgery, tumour classification and adjuvant treatment types were reviewed. These factors were poorly investigated across most studies. There were mixed findings for duration of mutism in relation to intellectual outcomes between two studies. In general, there was little variability in tumour classification and treatments across studies. Two studies found increased neurocognitive difficulties associated with greater time since surgery. However, the lack of consistent investigation of clinical factors limits certainty and generalisability of findings to aid prognosis.

### Limitations and directions for future research

Most studies included very small sample sizes of children with PPCMS, which were often compared to an unmatched, larger PPCMS-comparison group and increases the risk of falsely accepting the null hypothesis where groups were compared. Whilst small samples are common for rare conditions that affect survival, the small samples in the reviewed studies are exacerbated where identification of children with PPCMS was incidental to the research question or resulted in their exclusion from studies. Study findings are also limited by the absence of universally accepted diagnostic classification and screening for PPCMS. Many studies either did not provide clear criteria for PPCMS or described retrospective diagnosis. This produces implicit and hard to control variability across studies that may also contribute to variability in neurocognitive outcomes reported. PPCMS is unlikely to be a unitary construct with evidence that symptoms occur on a spectrum (2, 63) which may lead to underrepresentation of milder cases in the absence of systematic screening.

It is important to establish research programmes dedicated to the study of neurocognitive function after PPCMS, including research into potential measures for reducing neurocognitive sequelae and informing rehabilitative care. Provision of neurocognitive assessment should be prioritised in multicentre studies of PPCMS, preferably with consistent neurocognitive test batteries such as those following the European Society of Paediatric Oncology ‘core plus’ model for paediatric neuro-oncology (64). Both research and clinical practice will benefit from diagnostic clarity and routine prospective screening of PPCMS. Routine diagnostic neuropsychological assessments at key time points after posterior fossa tumour surgery for children will be supportive in both identifying those children most vulnerable to cognitive impairment and in need of support, and for treatment outcome monitoring. While there is ongoing research to develop a greater understanding of PPCMS, it is surprising how few studies have combined neurocognitive and neuroimaging findings which has great potential to further understanding of the pathophysiology and risk stratification of patients.

## 4.0 Conclusion

Despite limitations, this systematic review demonstrates evidence of increased vulnerability to neurocognitive impairment for children who experience PPCMS, even when compared to other children treated for posterior fossa tumour who are already an at-risk group. Vulnerabilities are particularly evident in executive functions, attention, processing speed, and psychomotor function which persist beyond the recovery of initial PPCMS symptoms in the post-operative phase and should inform rehabilitative care. It is hoped that the findings of this review better equip clinicians to plan assessments and interventions for children with PPCMS, inform general estimates of long-term prognosis following recovery of initial transient symptoms, and inform the systems supporting children affected by PPCMS on their long-term needs. There remains a need for dedicated research into the longitudinal trajectory of neurocognition in PPCMS and the impact of potentially moderating clinical factors such as duration and severity of mutism, age, and time since surgery on neurocognitive outcomes. Quality of life is poorer for children who experience PPCMS compared to those children who do not (65). Supporting affected families and children to develop an increased awareness and knowledge of their condition and long-term needs has helped to improve quality of life for children with PPCMS (66) and it is hoped that this will motivate clinicians and researchers to further our understanding of the condition.

### Involvement from People with Lived Experience

The systematic review was motivated and informed by the children and families we care for who have been affected by PPCMS. The Danny Green Fund, a charity that supports families affected by PPCMS, and the caregivers of children affected by PPCMS provided general guidance on the relevance and recommendations of the review. Their contributions specifically informed recommendations on 1) the need for routine screening for PPCMS symptoms using established diagnostic criteria, 2) the need to increase awareness of PPCMS and associated long-term effects within medical, familial, and educational contexts, and 3) the need for further research into neurocognitive outcomes in PPCMS.

## Data Availability

All data related to the present work are contained in the manuscript. Data sharing is not applicable to this article as no new data were created or analysed in this study.

## Abbreviations

PPCMS: Post-operative Paediatric Cerebellar Mutism Syndrome
PRISMA: Preferred Reporting Items for Systematic Reviews and Meta Analyses
MeSH: Medical Subject Headings
QUIPS: Quality in Prognostic Studies
IHE: Institute of Health Economics

## Acknowledgements

We thank the Danny Green Fund and the families of children affected by PPCMS at our institution for their invaluable contributions to this systematic review. All research at Great Ormond Street Hospital NHS Foundation Trust and UCL Great Ormond Street Institute of Child Health is made possible by the NIHR Great Ormond Street Hospital Biomedical Research Centre. The views expressed are those of the authors and not necessarily those of the NHS, the NIHR or the Department of Health.

## Author Contributions

**Conceptualisation:** BM Horne, K Aquilina, T Murphy, CP Malcolm. **Methodology**: BM Horne, T Murphy, CP Malcolm. **Data Acquisition, Analysis, and Interpretation**: BM Horne, AA Attanayake, T Murphy, CP Malcolm. **Validation**: BM Horne, AA Attanayake, T Murphy, CP Malcolm. **Visualisation:** BM Horne. **Supervision & Project Administration**: CP Malcolm & T Murphy. **Writing – Original Draft, & Writing – Review and Editing:** BM Horne, AA Attanayake, K Aquilina, T Murphy, CP Malcolm. All the authors have read the manuscript and agreed to its being submitted for publication.

## Declaration of competing interest

The authors have no conflicting interests to declare.

## Data Availability Statement

Data sharing is not applicable to this article as no new data were created or analysed in this study.

## Funding

None.

## Appendix

## Appendix S1

### Quality Appraisal

Risk of bias of included studies.

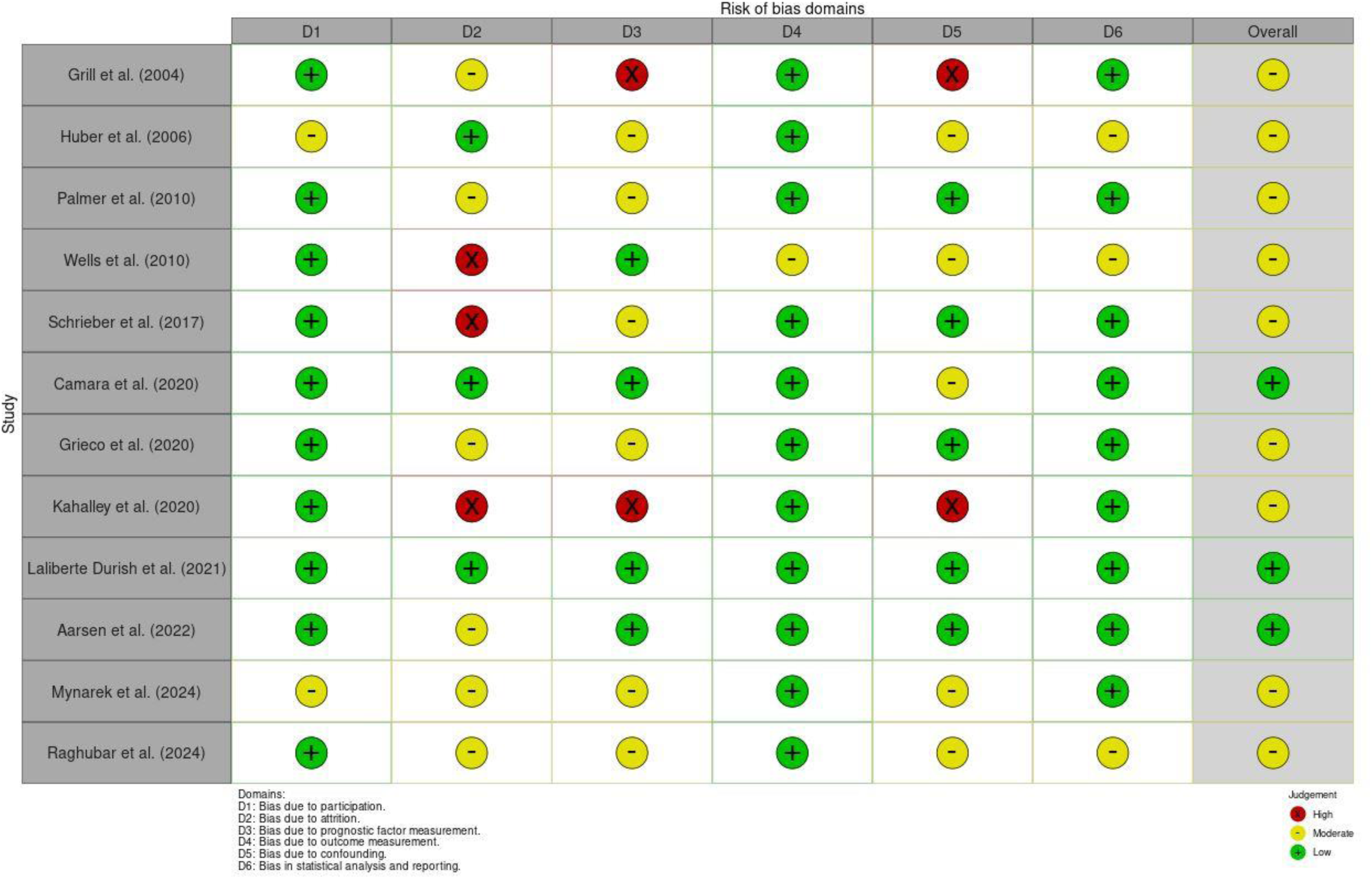
Risk of bias in included cohort studies assessed using the QUIPS tool.

|  | Risk of bias domains |  |  |  |  |  | Overall |
| --- | --- | --- | --- | --- | --- | --- | --- |
|  | D1 | D2 | D3 | D4 | D5 | D6 |  |
| Grill et al. (2004) | + | - | X | + | X | + | - |
| Huber et al. (2006) | - | + | - | + | - | - | - |
| Palmer et al. (2010) | + | - | - | + | + | + | - |
| Wells et al. (2010) | + | X | + | - | - | - | - |
| Schrieber et al. (2017) | + | X | - | + | + | + | - |
| Camara et al. (2020) | + | + | + | + | - | + | + |
| Grieco et al. (2020) | + | - | - | + | + | + | - |
| Kahalley et al. (2020) | + | X | X | + | X | + | - |
| Laliberte Durish et al. (2021) | + | + | + | + | + | + | + |
| Aarsen et al. (2022) | + | - | + | + | + | + | + |
| Mynarek et al. (2024) | - | - | - | + | - | + | - |
| Raghubar et al. (2024) | + | - | - | + | - | - | - |

Study
Domains: D1: Bias due to participation. D2: Bias due to attrition. D3: Bias due to prognostic factor measurement. D4: Bias due to outcome measurement. D5: Bias due to confounding. D6: Bias in statistical analysis and reporting.
Judgement High Moderate Low

Risk of bias in included case series assessed using the IHE tool.

| Study | Risk of bias |  |  |  |  |  |  |  |  |  |  |  |  |  |  |  |  |  |  |  | Overall |
| --- | --- | --- | --- | --- | --- | --- | --- | --- | --- | --- | --- | --- | --- | --- | --- | --- | --- | --- | --- | --- | --- |
|  | D1 | D2 | D3 | D4 | D5 | D6 | D7 | D8 | D9 | D10 | D11 | D12 | D13 | D14 | D15 | D16 | D17 | D18 | D19 | D20 |  |
| De Smet et al. (2009) | + | - | X | X | + | + | + | + | + | + | X | + | X | + | + | + | + | + | + | X | - |
| Di Rocco et al (2011) | + | + | X | X | + | + | + | + | + | + | X | + | + | + | - | - | + | + | + | + | - |
| Kupeli et al. (2011) | + | - | - | X | + | + | + | + | - | + | X | + | X | + | - | + | X | + | + | X | - |
| Fay McClymont et al. (2017) | + | X | + | X | X | - | - | X | X | + | X | + | X | + | + | - | + | X | + | X | - |

D1: Was the hypothesis/aim/objective of the study clearly stated? D2: Was the study conducted prospectively? D3: Were the cases collected in more than one centre? D4: Were patients recruited consecutively? D5: Were the characteristics of the patients included in the study described? D6: Were the eligibility criteria (i.e. inclusion and exclusion criteria) for entry into the study clearly stated? D7: Did patients enter the study at a similar point in the disease? D8: Was the intervention of interest clearly described? D9: Were additional interventions (co-interventions) clearly described? D10: Were relevant outcome measures established a priori? D11: Were outcome assessors blinded to the intervention that patients received? D12: Were the relevant outcomes measured using appropriate objective/subjective methods? D13: Were the relevant outcome measures made before and after the intervention? D14: Were the statistical tests used to assess the relevant outcomes appropriate? D15: Was follow-up long enough for important events and outcomes to occur? D16: Were losses to follow-up reported? D17: Did the study provide estimates of random variability in the data analysis of relevant outcomes? D18: Were the adverse events reported? D19: Were the conclusions of the study supported by the results? D20: Were both competing interests and sources of support for the study reported?
Judgement X High - Unclear + Low

## Notes

### Competing Interest Statement

The authors have declared no competing interest.

### Funding Statement

This study did not receive any funding

